# Pediatric intensive care unit admissions for COVID-19: insights using state-level data

**DOI:** 10.1101/2020.07.22.20160226

**Authors:** Rohit S. Loomba, Enrique G. Villarreal, Juan S. Farias, Ronald A. Bronicki, Saul Flores

## Abstract

**Introduction:** Intensive care has played a pivotal role during the COVID-19 pandemic as many patients developed severe pulmonary complications. The availability of information in pediatric intensive care (PICUs) remains limited. The purpose of this study is to characterize COVID-19 positive admissions (CPAs) in the United States and to determine factors that may impact those admissions.

**Materials and Methods:** This is a retrospective cohort study using data from the COVID-19 dashboard virtual pediatric system containing information regarding respiratory support and comorbidities for all CPAs between March and April 2020. The state level data contained 13 different factors from population density, comorbid conditions and social distancing score. The absolute CPAs count was converted to frequency using the state’s population. Univariate and multivariate regression analyses were performed to assess the association between CPAs frequency and endpoints.

**Results:** A total of 205 CPAs were reported by 167 PICUs across 48 states. The estimated CPAs frequency was 2.8 per million children. A total of 3,235 tests were conducted with 6.3% positive tests. Children above 11 years of age comprised 69.7% of the total cohort and 35.1% had moderated or severe comorbidities. The median duration of a CPA was 4.9 days [1.25-12.00 days]. Out of the 1,132 total CPA days, 592 [52.2%] were for mechanical ventilation. The inpatient mortalities were 3 [1.4%]. Multivariate analyses demonstrated an association between CPAs with greater population density [beta-coefficient 0.01, p<0.01] and increased percent of children receiving the influenza vaccination [beta-coefficient 0.17, p=0.01].

**Conclusions:** Inpatient mortality during PICU CPAs is relatively low at 1.4%. CPA frequency seems to be impacted by population density while characteristics of illness severity appear to be associated with ultraviolet index, temperature, and comorbidities such as Type 1 diabetes. These factors should be included in future studies using patient-level data.

## Introduction

During the first quarter of 2020, the world has been subdued by the rapid worldwide spread of the novel Severe Acute Respiratory Syndrome Coronavirus (SARS-CoV-2). On March 11th, 2020 the World Health Organization declared the disease caused by the novel virus (COVID-19) a pandemic health emergency for the first time since the swine flu (H1N1) in 2009. As of May 2020, there were more than 3.2 million people infected, with more than 670,000 deaths worldwide. The United States (US) has been one of the most impacted nations with more than 1 million people infected and more than 60,000 deaths as of May 1st, 2020[1]. Hospitals in areas where the pandemic has caused devastation continue to struggle as many challenges remain unmet due to the speed of transmission, the lack of accurate knowledge regarding the benefits or pitfalls of the current available therapies and the uncertainty of being able to provide adequate care if the rate of transmission continues.

The availability of intensive level of care has played a pivotal role, as many patients developed severe pulmonary complications. However, there is limited information on COVID-19 positive admissions (CPAs) to pediatric intensive care units (PICUs) regarding patient characteristics, respiratory support required and their impact on outcomes. Therefore, the purpose of this study is to better characterize CPAs to PICUs in the US and to determine factors that may impact those admissions.

## Methods

This study utilized only publicly available, deidentified, state-level data. As such, no institutional review board review or approval was sought.

### Endpoint identification and data collection

The following data was identified for collection regarding the CPAs themselves: number, duration, need for various ventilatory support measures, severity of comorbidities, and the total number of COVID-19 tests conducted. The following data was collected regarding US states: pediatric population, state population (pediatric and adult) density, air and drinking water quality, average temperature, average ultraviolet index, prevalence of pediatric obesity, type 1 diabetes mellitus (DM) and asthma, the proportion of children who smoke cigarettes, received the influenza vaccine, had health insurance, and received home health care, race, percent of households with children below the poverty line, highest education level of adults in homes with children, and the social distancing score by global positional satellite data (Supplementary Table 1).

The data regarding the CPAs themselves was collected from the publicly available COVID-19 dashboard provided by the Virtual Pediatric System (VPS), which collects data from several PICUs in the US. COVID-19 data was collected from March 14^th^ through April 14^th^, 2020, in order to represent one full month of data. Data regarding number of centers, number of tests, and number of CPAs was captured in absolute counts. Data regarding CPAs duration was collected in days. The respiratory support modalities for which data was available were room air (RA), nasal cannula (NC) and for the advanced respiratory support modalities (i.e. other than RA and NC) there was available data for high flow nasal cannula (HFNC), non-invasive positive pressure ventilation (NIPPV), conventional mechanical ventilation (MCV), high frequency oscillatory ventilation (HFOV), and extracorporeal membrane oxygenation (ECMO), and was captured in duration (days) of their use. Data regarding severity of comorbidities is reported in the VPS dashboard and the percentage of CPAs with moderate or severe degree of comorbidities was collected.

State-wide data for the analyses were collected from a variety of sources with the complete list of sources provided as Supplementary Material 1. Children’s population data and pediatric comorbidity data was obtained from 2018, as these were the most recent and comprehensive data available. The sources for these other data points were generally US government-based efforts to capture state-level data on various medical issues, however, not all states reported data for all the endpoints (Supplementary Table 2).

Endpoints were assigned to the authors for collection. One author was responsible for collecting data for each state for the variables assigned. Once these data were collected a different author, who did not primarily collect data for that specific endpoint, verified the numbers for accuracy. Finally, values in the top and bottom 10^th^ percentile were identified and verified by a third author.

### Statistical analyses

As the data was collected for each state and intended for state-level analyses, and each state has a different pediatric population, the absolute numbers of CPAs for each state were not directly comparable. Thus, the absolute CPAs count for each state was first converted to a frequency of CPAs per 1,000,000 children using the specific state’s population. This CPAs frequency was then used as the dependent variable in a series of single-independent variable linear regressions to determine the univariate association between CPAs frequency and the other endpoints. Multivariate regression was conducted with CPAs frequency as the dependent variable and with other variables entered as independent variables. Forward stepwise regression was utilized with the model with greatest R-squared value being used for the analyses.

Next, a composite endpoint called “percent of PICUs days requiring advanced respiratory support” was created. This consisted of the total duration of HFNC, NIPPV, MCV, HFOV, and ECMO divided by the total PICUs admission duration. This was then modeled similarly to CPAs frequency. Next, a composite outcome called “percent of PICU days requiring intubation” was created. This consisted of the total duration of MCV and HFOV divided by the total PICU admission duration. This, too, was then modeled similarly as CPA frequency. Lastly, an endpoint called “PICUs duration per admission” was created for each state and consisted of the total CPAs PICUs duration for that specific state divided by the number of CPAs reported by that state. This was also then modeled similarly to CPA frequency.

All statistical analyses were done using the user-coded, syntax-based interface of SPSS Version 23.0. A p-value of 0.05 was considered statistically significant. All statistical analyses were done at the state-level with state-level data. Analyses were not conducted at a patient-level with patient-level data. Any use of the word significant here-on in the manuscript refers to “statistically significant” unless explicitly specified otherwise.

## *Re*sults

### COVID-19 pediatric intensive care unit admission characteristics

A total of 205 CPAs were reported by 167 PICUs across 48 states. The states not represented in the VPS dataset were Maine and Wyoming. By using an estimated pediatric population of the US of 72,886,669, the frequency of CPAs was 2.8 per million children (Figure 1a). The median CPAs frequency for a state was 0.66 with a range of 0 to 18.68. A total of 3,235 tests were conducted by all reporting sites, resulting in 6.3% of total tests resulting positive (Table 1).

**Figure 1.**
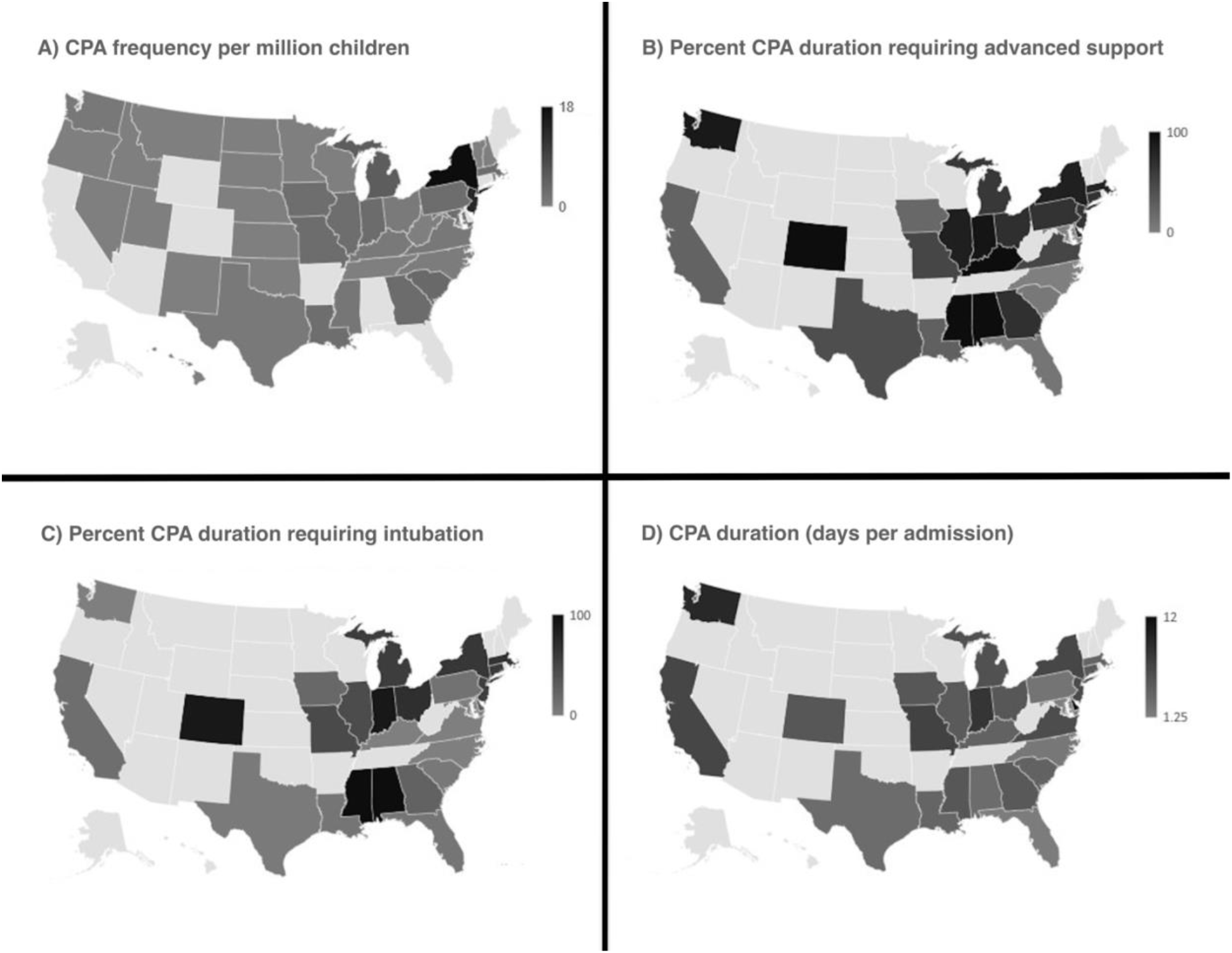
COVID-19 pediatric intensive care unit admission characteristics by state. A) COVID-19 pediatric admission frequency per million children. B) Percent of COVID-19 pediatric admission duration requiring advanced respiratory support. C) Percent of COVID-19 pediatric admission requiring intubation. D) COVID-19 pediatric admission duration: days per admission. CPA indicates COVID-19 pediatric admission.

**Table 1.**
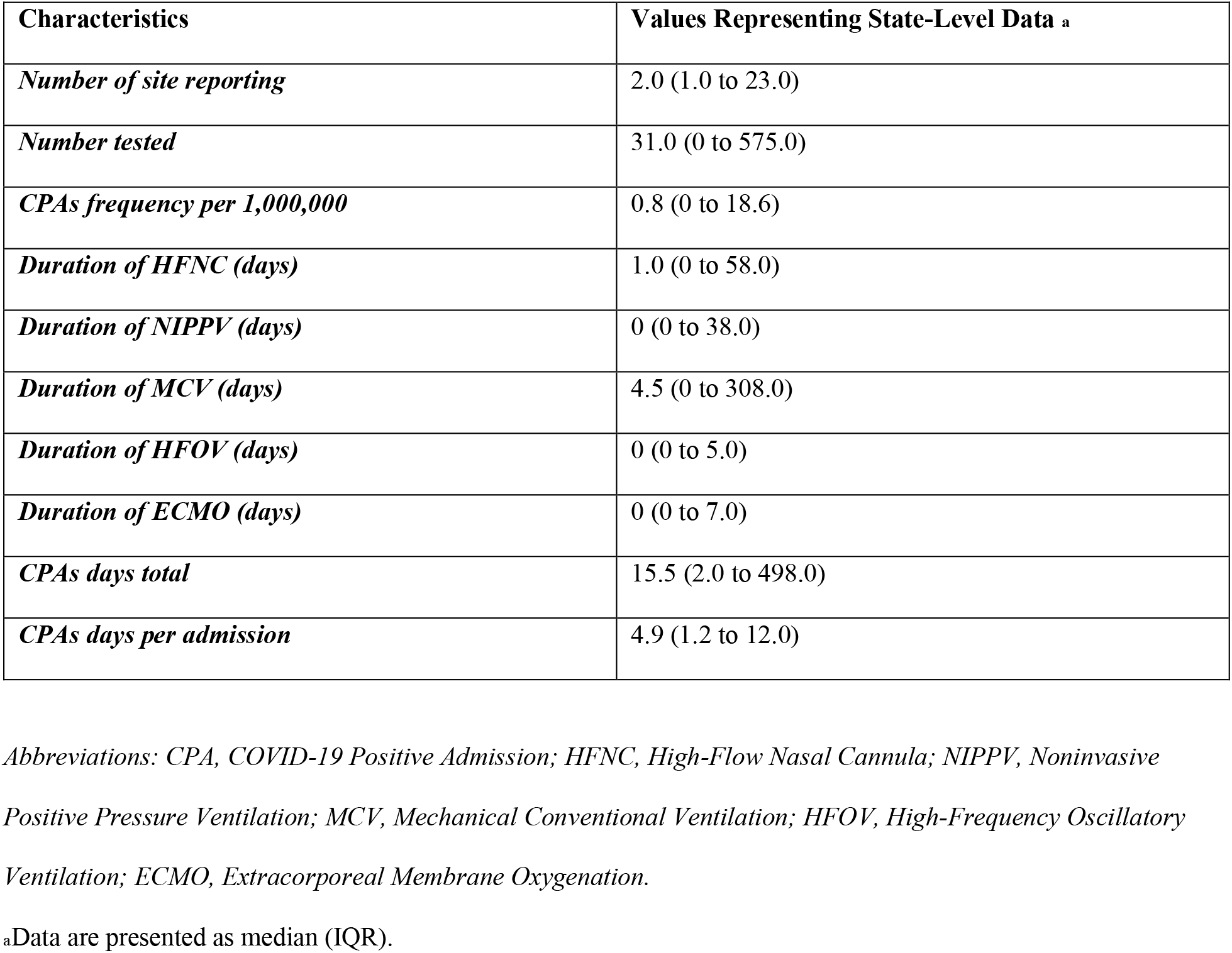
Demographic Characteristics of Children Treated in Pediatric Intensive Care Units for COVID-19 in the United States of America.

Of the 205 CPAs, 31 (15.1%) were for children 2 years of age or under, while 32 (15.6%) were for children between 2 and 11 years of age, and the remainder (69.7%) above 11 years of age. Of the 205 CPAs, 70 (33.6%) had mild severity of comorbidities while 73 (35.1%) had moderate or severe comorbidities.

The total CPAs duration for all 205 CPAs was 1,132 days. Out of the 1,132 CPAs days, 309 days (27.3%) were on nasal cannula or room air, 129 (11.6%) consisted of HFNC, 83 (7.3%) of NIPPV, 592 (52.2%) of MCV, 6 of HFOV, and 13 (1.6%) of ECMO (Figure 1b and 1c). The median duration of a CPAs was 4.9 days, with a range of 1.25 to 12.00 days (Figure 1d).

Of the 205 CPAs, 3 (1.4%) ended in inpatient mortality.

### COVID-19 pediatric intensive care unit admission frequency, univariate analyses

Univariate analyses with CPAs frequency as the dependent variable demonstrated the following factors to be significantly associated with increasing CPAs frequency: greater population density (beta-coefficient 0.01, p< 0.01) and increased percent of children receiving the influenza vaccination (beta-coefficient 0.17, p=0.01). None of the independent variables were associated with significant reductions in CPAs frequency (Table 2).

**Table 2.** Univariate Analysis: Association between pediatric population characteristics and COVID-19 positive admissions.

|  | CPA frequency |  | Percent CPA duration<br>requiring HFNC or<br>more |  | Percent CPA duration<br>requiring intubation<br>(MCV or HFOV) |  | Number of PICU days<br>per CPA |  |
| --- | --- | --- | --- | --- | --- | --- | --- | --- |
|  | Beta-coefficient | P-value<sub>a</sub> | Beta-coefficient | P-value<sub>a</sub> | Beta-coefficient | P-value<sub>a</sub> | Beta-coefficient | P-value<sub>a</sub> |
| *Population density* | 0.008 | 0.004 | -0.011 | 0.685 | 0.011 | 0.647 | -0.00008 | 0.996 |
| *Urban air quality* | 0.032 | 0.404 | -0.209 | 0.305 | 0.098 | 0.856 | 0.07 | 0.864 |
| *Drinking water quality* | -0.055 | 0.147 | -0.207 | 0.765 | -0.309 | 0.636 | 0.032 | 0.48 |
| *UV index* | -0.614 | 0.182 | -12.008 | 0.04 | -5.821 | 0.308 | -0.782 | 0.044 |
| *Average precipitation* | 0.421 | 0.185 | 8.94 | 0.084 | 6.308 | 0.203 | 0.116 | 0.743 |
| *Average temperature* | 0.018 | 0.708 | -1.296 | 0.064 | -0.768 | 0.256 | -0.093 | 0.045 |
| *Percent households below poverty line* | 0.053 | 0.612 | -0.399 | 0.784 | 0.348 | 0.801 | -0.083 | 0.39 |
| *Percent households with below high school as*<br>*highest adult education level* | 0.111 | 0.502 | -3.926 | 0.166 | -3.182 | 0.238 | -0.032 | 0.866 |
| *Percent pediatric obesity* | 0.067 | 0.614 | -2.187 | 0.28 | -0.391 | 0.84 | -0.179 | 0.179 |
| *Percent pediatric type 1 DM* | -0.052 | 0.68 | 4.525 | 0.012 | 4.899 | 0.003 | 0.116 | 0.356 |
| *Percent pediatric asthma* | 0.175 | 0.779 | -3.707 | 0.564 | 1.534 | 0.821 | -0.079 | 0.851 |
| *Percent high schoolers currently smoking* | -1.48 | 0.524 | -0.726 | 0.836 | -3.397 | 0.3 | -0.269 | 0.242 |
| *Percent pediatric flu shot received* | 0.177 | 0.019 | 0.252 | 0.814 | -0.22 | 0.828 | 0.026 | 0.72 |
| *Percent pediatric who have health insurance* | 0.24 | 0.294 | 2.368 | 0.438 | 2.604 | 0.366 | 0.225 | 0.264 |
| *Percent pediatric who have a medical home* | -0.098 | 0.449 | 1.486 | 0.425 | 0.715 | 0.687 | 0.066 | 0.595 |
| *Percent hispanic* | 0.015 | 0.725 | -0.37 | 0.535 | -0.468 | 0.405 | 0.001 | 0.99 |
| *Percent white* | -0.038 | 0.256 | 0.799 | 0.149 | 0.587 | 0.266 | 0.02 | 0.589 |
| *Percent black* | 0.093 | 0.072 | -0.741 | 0.279 | 0.113 | 0.863 | -0.05 | 0.27 |
| *Percent other race* | -0.027 | 0.638 | 0.376 | 0.848 | -2.903 | 0.109 | 0.156 | 0.224 |
| *Social distancing score* | 0.178 | 0.848 | 17.57 | 0.196 | 3.558 | 0.786 | 1.664 | 0.061 |
| *Moderate or severe comorbidity<sup>b</sup>* |  |  | 0.088 | 0.686 | -0.009 | 0.965 | 0.014 | 0.33 |

<sup>a</sup> P-value <0.05 was considered statistically significant.
<sup>b</sup> No data available to assess moderate or severe comorbidity with admission frequency.

### Covid-19 pediatric intensive care unit admission frequency, multivariate analyses

Multivariate regression analysis demonstrated the following to be associated with increased CPAs frequency: population density (beta coefficient 0.1, p< 0.01). The R-square value was 0.28. There was no collinearity present (Table 3).

**Table 3.**
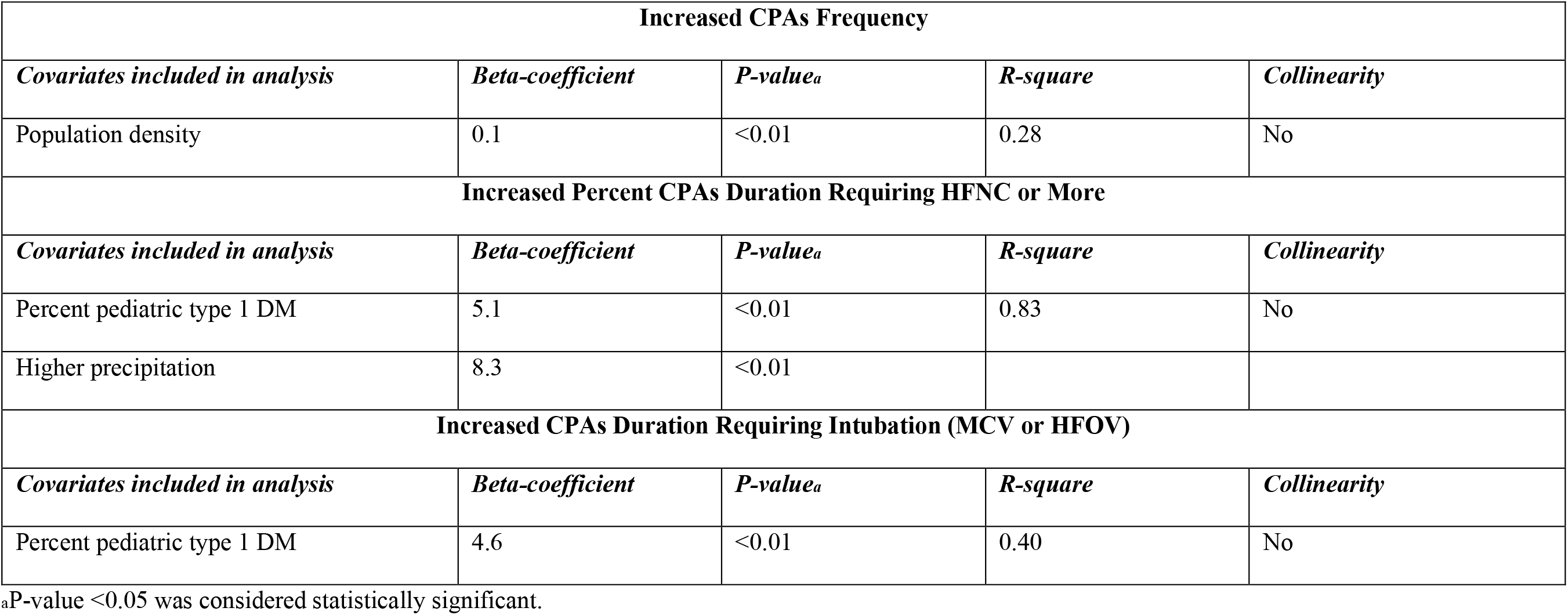
Multivariate Regression Analyses.

| **Increased CPAs Frequency** |  |  |  |  |
| --- | --- | --- | --- | --- |
| *Covariates included in analysis* | *Beta-coefficient* | *P-value<sub>a</sub>* | *R-square* | *Collinearity* |
| Population density | 0.1 | <0.01 | 0.28 | No |
| **Increased Percent CPAs Duration Requiring HFNC or More** |  |  |  |  |
| *Covariates included in analysis* | *Beta-coefficient* | *P-value<sub>a</sub>* | *R-square* | *Collinearity* |
| Percent pediatric type 1 DM | 5.1 | <0.01 | 0.83 | No |
| Higher precipitation | 8.3 | <0.01 |  |  |
| **Increased CPAs Duration Requiring Intubation (MCV or HFOV)** |  |  |  |  |
| *Covariates included in analysis* | *Beta-coefficient* | *P-value<sub>a</sub>* | *R-square* | *Collinearity* |
| Percent pediatric type 1 DM | 4.6 | <0.01 | 0.40 | No |

<sup>a</sup>P-value <0.05 was considered statistically significant.

### COVID-19 pediatric intensive care unit admission requirement of advanced respiratory support, univariate analyses

Univariate analyses with the percent of CPAs duration requiring advanced respiratory support as the dependent variable demonstrated the following factors to be associated with a significant increase in the duration: increased prevalence of type 1 DM (beta-coefficient 4.52, p=0.01). The following were associated with a decrease in this duration: ultraviolet light index (beta-coefficient −12.00, p=0.04) (Table 2).

### Covid-19 pediatric intensive care unit admission requirement of advanced respiratory support, multivariate analyses

Multivariate regression analysis demonstrated the following to be associated with increased percentage of CPAs duration requiring advanced respiratory support: higher prevalence of type 1 diabetes (beta coefficient 5.1, p< 0.01) and higher precipitation (beta coefficient 8.3, p< 0.01). The R-square value was 0.83. There was no collinearity present (Table 3).

### COVID-19 pediatric intensive care unit admission requirement of intubation, univariate analyses

Univariate analyses with the percent of CPAs duration requiring intubation as the dependent variable demonstrated the following factors to be associated with a significant increase in the duration: increased prevalence of type 1 DM (beta-coefficient 4.89, p<0.01). There were no factors associated with a significant decrease in percent of CPAs duration requiring intubation (Table 2).

### Covid-19 pediatric intensive care unit admission requirement of intubation, multivariate analyses

Multivariate regression analysis demonstrated the following to be associated with increased percentage of CPAs duration requiring intubation: prevalence of type 1 diabetes (beta coefficient 4.6, p< 0.01). The R-square value was 0.40. There was no collinearity present (Table 3).

### COVID-19 pediatric intensive care unit admission duration, univariate analyses

Univariate analyses with CPAs duration as the dependent variable demonstrated the following factors to be significantly associated with decreased admission duration: increased ultraviolet light index (beta-coefficient −0.78, p=0.04), increased average temperature (−0.09, p=0.04). There were no factors associated with a significant increase in CPAs duration (Table 2).

### Covid-19 pediatric intensive care unit admission duration, multivariate analyses

None of the variables remained statistically significant in multivariate analysis for CPA duration. Thus, no model is presented.

### Power analyses

Multivariate regression analysis for CPAs frequency, with its low effect size and one independent variable, would require 385 subjects to achieve 80% power. Thus, this model was not adequately powered.

Multivariate regression analysis for percentage of CPAs duration requiring advanced respiratory support, with its low effect size and one independent variable, would require 31 subjects to achieve 80% power. Thus, this model was adequately powered.

Multivariate regression analysis for percentage of CPAs duration requiring intubation, with its moderate effect size, would require 49 subjects to achieve 80% power. Thus, this model was adequately powered.

No variables were of substantial enough significance for a multivariate regression for CPAs admission duration.

## Discussion

These analyses identified several important factors in PICUs CPAs in the US. First, the burden of COVID-19 in children has not been nearly as substantial as in the adults, with a CPAs frequency of 2.8 per million children. The highest 5-day rolling census average noted in the VPS dashboard was 90 CPAs. If this number is doubled to account for PICUs not reporting to VPS, that would imply a maximum CPAs burden of 180 in a single day. Considering that there are approximately 5,100 PICUs beds in the US and a down trending daily PICUs census and new CPAs numbers reported in the VPS dashboard, it is unlikely to become a burden on the system during the study time. This is an important finding as a recent model reported a possibility of a higher burden of disease without providing a time frame for cases to present[2]. Second, these analyses demonstrate that CPAs have a relatively low mortality rate of 1.4%. Third, slightly over half of CPAs required intubation. Fourth, the median CPAs duration over the study period was 4.9 days.

These findings are consistent with current national and international trends. In the US, the Morbidity and Mortality Weekly Report stated that pediatric cases represent 1.7% of the total cases[3], whereas in Italy and China, the cases represented 1.2% and 2%, respectively [4, 5]. Furthermore, the majority of pediatric patients presented with mild symptoms having a complete recovery within 1 to 2 weeks after the onset of illness [6]. A study by Dong and colleagues reported in a cohort of 2,135 confirmed and suspected pediatric COVID-19 cases, 5.8% were severe or critical and there was 1 mortality[7].

The association with state population density makes intuitive sense as urban centers potentiate a rapid spread of an infectious disease, as well as a greater number of global travel routes[8]. Though, it was reported that influenza immunization rates are higher in urban areas[9], the role of influenza immunization in the COVID-19 transmission requires further investigation as recent studies have described the phenomenon of virus interference in which an immunization against one virus may increase the risk of illness from other viruses[10]. A recent study by Wolff and colleagues found in a cohort of 6,000 patients, influenza immunization increased the risk of illness from non-pandemic coronavirus (odds ratio of 1.36; 95% confidence interval 1.14 to 1.63, p< 0.01)[11]. Similarly, Rikin and colleagues identified an increased risk of non-influenza respiratory illness in children who received the influenza vaccine when compared to those who did not (hazard ratio 1.65, 95% confidence interval 1.14 to 2.38). Future studies on pediatric COVID-19 should include influenza vaccination as one of the variables of interest. There is limited data on the association between environmental temperature or ultraviolet index with the burden of COVID-19 in children. Our findings showed an association between an increase in ultraviolet index and temperature with a reduction in duration of advanced respiratory support (i.e. increased from room air or nasal cannula). An association was identified by Gunthe and colleagues with lower environmental temperatures between 5 to 15 degree Celsius (41 to 59 degree Fahrenheit) and a low ultraviolet index of 2.5 with an increased number of COVID-19 cases[12]. This findings are in alignment with prior studies on SARS-CoV-1 where high temperature and high humidity decreased its survival on contaminated surfaces[13]. Further studies are warranted to determine the implications of environmental factors in SARS-CoV-2 life cycle.

Our study findings showed an association between PICU CPAs requiring intubations and prevalence of type 1 DM. A study by Shekerdemian and colleagues identified in a cohort of 48 confirmed COVID-19 critically ill pediatric cases, 4 (8%) with type 1 DM with 3 of them presenting with diabetic ketoacidosis[14]. Future studies should focus on determining the role of DM in the pathogenesis and severity of COVID-19 in children.

As the mechanisms of viral spread remain largely undefined, countries have engaged in the implementation of social distancing as the main preventive measure. However, the role of children in spreading SARS-CoV-2 is highly speculated and presents conflicting answers with some reports stating similar virus spreading between children and adults[15], where as other reports from Wuhan and Shanghai state that children are less likely than adults to spread the virus[16]. Some European nations like Sweden have faced the current pandemic with alternate strategies including more lenient social distancing efforts and have not had, to date, severely adverse outcomes as a result in children[17]. The results of our analysis may support the latter since children represent a small fraction of confirmed COVID-19 cases. There is limited information to determine the impact of social distancing and lockdowns during this pandemic, previously published modeling data demonstrates that, in certain circumstances, social distancing may have no impact on the illness, and in certain circumstances may have negative population outcomes [18]. Studies of transmission in schools will be important as schools get ready to reopen. Meanwhile, the population should follow the recommendations from local municipalities.

These analyses offer an early-look at COVID-19 PICUs admissions and offer some descriptive insight into these admissions as well as use state-wide data to investigate associations between CPAs characteristics with comorbidities, environmental factors, and socioeconomic factors. While these analyses offer novel data regarding CPAs in the US, they are not without their limitations. Firstly, all the data is state-wide data as no patient-level data was used. Thus, the 48 states with reported data are the cases, not the 205 individual CPAs. The data regarding comorbidities, environmental factors, and socioeconomic factors is also state-level data and is not specific to the CPAs. Rather, the information used is publicly available data about the children or families in the respective state. Additionally, the multivariate analyses are underpowered and thus the univariate analyses in this study offer the more meaningful data. It must be kept in mind that all significant findings in this study are simply associations as causation cannot be inferred due to the study design. Thus, findings such as those related to immunizations and social distancing, cannot be considered to be causal and should not be utilized to impact decision-making.

Despite the limitations outlined, these analyses offer helpful information that may be used to assist during the consideration as to what factors need more clarification from future studies with patient-level data. These results also offer a cross-sectional view regarding US PICU CPAs not previously reported. The use of state-level data allowed for analyses relatively early, before aggregate, multicenter, patient-level data-based studies can be completed, thus, it can used while the pandemic is still occurring. Furthermore, these analyses also offer insight into some of the factors that should be included in future studies and may also be helpful for resource allocation planning and CPAs admission.

## Conclusion

Inpatient mortality during PICU CPAs is relatively low at 1.4%. CPA frequency seems to be impacted by population density while characteristics of illness severity appear to be associated with ultraviolet index, temperature, and comorbidities such as Type 1 diabetes. T these factors should be included in future studies using patient-level data.

## Data Availability

Deidentified individual participant data will be made available at https://figshare.com/articles/dataset/Pediatric_intensive_care_unit_admissions_for_COVID-19_insights_using_state-level_data/12693461

## Acknowledgments

None.

## Supplementary material

Supplementary Table 1. Information Resource for state-wide data collection

Supplementary Table 2. States that reported data for each outcome

